# Prevalence of Gender-based Violence and Factors Affecting Help Seeking Among Females Aged 15 Years and Above in Ayilo Refugee Camp in Adjumani District, Uganda

**DOI:** 10.1101/2025.09.08.25335372

**Authors:** Denish Odongo, Michael Job Aeku, Boniface Ocora, Jerom Okot, Kenneth Opiro, Pebalo Pebolo Francis

## Abstract

**Background:** Gender-based violence is one of the most pervasive and hideous form of human rights violations that demeans the very meaning of existence of various groups based on their different gender and roles. It is worsened in emergency situations where social support structures have broken down. It is estimated that the protracted South Sudan crisis has caused devastation with over 5 million people in dire need of humanitarian aid including 1.2 million women and girls of reproductive age. Despite the available policies and well-established care pathways, there is a notably vivid trend of underutilization and under reporting of cases. This study was therefore to find out the exact prevalence of GBV and the factors affecting help seeking among women aged 15 years and above in Ayilo refugee camp.

**Methods:** A cross-sectional study was done using both Key informant interviews and semi-structured questionnaires administered to participants. Data was analyzed using Microsoft Excel, descriptive statistics were presented in tables, charts, and narratives. Secondary reentry was done using kobo collect and exported to STATA for statistical analysis at bivariate and multivariate levels.

**Results:** The prevalence of GBV was at 54% with a likelihood that it is even higher because of under reporting. Emotional and physical violence was the most prevalent form of violence. Only about half of the survivors sought redress, and mainly from the family and social networks compared with other sectors in the victim care pathway. Factors affecting help seeking included stigma and fear of being exposed, suppression by culture and cultural practices especially bride price, lack of confidentiality from the people in the care pathway, consequences of reporting and influence of local leaders on most cases.

**Conclusion:** The prevalence of violence is higher than reported. Help seeking was low due to survivor related factors, and socio-cultural influences despite existing victim care pathway. There is need to advocate for attitude and behavior change, improving services rendered and addressing socio-economic challenges faced by those at risk.

## Background

Gender-based violence (GBV) remains one of the most severe threats to women’s health and well-being, primarily driven by systemic inequity and social injustice(1,2). Rooted in unequal power dynamics between men and women, GBV inflicts various forms of harm sexual, physical, mental, behavioural, and economic particularly affecting women in both public and private spaces(3). Globally, over a third of women face some form of physical or sexual violence during their lives, with substantial negative impacts on reproductive, maternal, adolescent, and mental health(2,4).

The prevalence of GBV is notably high in sub-Saharan Africa, especially in conflict and post-conflict areas(5). For example, in Northern Uganda, over 80% of women have reported verbal or psychological abuse, and up to 23% have experienced sexual violence(6). Additionally, about 25% of women aged 15 years and older report experiencing violence in their lifetime. Women and young girls in refugee settlements are particularly vulnerable due to conflicts, insecurity, violence, and poverty(7).

Uganda’s open-door refugee policy has led to the influx of over 1.5 million refugees, predominantly from South Sudan and the Democratic Republic of Congo, with women and children comprising 81% of this population(6). The refugee settlements face significant challenges, including pervasive poverty, limited access to education and healthcare, and cultural norms surrounding gender and sexuality, all of which contribute to the prevalence of sexual and gender-based violence (SGBV) in these settings(8).

The West Nile region, which has the highest concentration of refugee camps in Uganda, hosts a refugee population of 626,331 and reports some of the highest rates of teenage pregnancies(9). The Ayilo refugee settlement, home mainly to Dinka and Madi ethnic groups, is characterized by cultural practices that elevate the male gender and may perpetuate GBV(10). Cultural norms, social constraints, and fear of stigmatization often deter women from seeking help after experiencing violence. As a result, the true extent of SGBV remains underreported, exacerbating the harmful effects such as sexually transmitted infections, fistulas, unwanted pregnancies from rape, physical injuries, stigmatization, substance abuse, and economic hardship(11).

Understanding the actual burden of GBV and identifying predictors of help-seeking behaviour among victims in refugee settings is crucial. This knowledge will enable targeted intervention strategies by implementing partners and the government to address and mitigate the consequences of GBV in these vulnerable populations. This study therefore determined the burden of GBV and the factors influencing help-seeking behaviour among victims in refugee settings in Uganda.

## Methods

### Design and Settings

This cross-sectional study, in which both quantitative and qualitative methods was carried out in Ayilo Refugee camp (1 and 2) located in Pakele Sub-County in Adjumani District, West Nile Sub-region of Uganda. Ayilo refugee camps host over 185,000 refugees.

### Study population and Sample size

Our study targeted female aged 15 years or older living in the refugee settlements of Northern Uganda. We included only respondents between 15 or older, who consented to our study, and those who did not consent to the study were not included. Fifteen years lower age limit was deliberately chosen as the age in which girl child starts puberty and develops feminine physiques that is culturally considered grown up by the community. In addition to these participants, four key informants from legal, medical, Camp leadership and law enforcement were interviewed. Only those who were sampled and were willing to participate in the study were enrolled. The males were excluded because in the South Sudan culture they are considered more superior and less likely to suffer GBV hence the study focused on the vulnerable female populations. The study excluded those who were still traumatized significantly, in health facilities or detention centers and/or identified by the care givers as recent victims. Those who asked for material and or monetary benefits following participation in the study were also excluded.

### Sampling and sample size

Using Kish and Leslie formula (1965) at 95% confidence intervals and 35% global prevalence according to the global and scientific review done by WHO in 2013 (Global and Regional Estimates of Violence Against Women, 2013), a total of 300 sample size was calculated. Four key informants were also interviewed.

Ayilo Refugee camp was randomly selected among the refugee camps in Northern Uganda. It is subdivided in to Ayilo 1 and 2 having 6 and 7 blocks respectively. Cluster sampling was used where Ayilo 1 and 2 formed clusters and the 13 blocks formed sub-clusters. Cluster 1 had 3531 households with a total population of 25,579. Cluster 2 had 2,215 households with a total population of 14,430.

A systematic sampling technique was then used to sample every 21^st^ and 17^th^ household which realized a total of 187 and 107 households respectively in cluster 1 and 2, from which only one female was interviewed per household selected by simple random sampling. This gave an average of 32 and 15 study units per block in cluster 1 and 2 respectively, and a total of 300 participants. Convenience sampling was used to identify the 4 key informants from the different GBV care pathways including a health GBV coordinator (reproductive health officer from MTI), a law enforcer (Police officer in charge of GBV), a community leader (the Camp block welfare officer) and the camp commandant who is the overall coordinator.

### Study Variables

The study variables included independent variables such as socio-demographic characteristics of the study population. While the dependent variable was history of GBV. GBV was defined as an abuse of power that inflicts harm on the survivor such as physical, emotional, or sexual in nature involving rape, physical assault, sexual abuse, or intimate partner violence. Participants were asked if they could remember been exposed to any of the above acts.

### Data Collection

Data were collected using a pretested researcher administered questionnaire with the help of translators (where applicable). The questionnaires were pretested a week earlier, and credibility and ease of use evaluated, and adjustments made before the study was conducted. Translators who could speak the predominant language in Ayilo settlement such as Madi, Kuku, Kakwa, Kiswahili and Dinka, and mainly either Village Health Teams (VHT) or GBV incentive care workers were used because they had training or understanding on GBV. They translated the questions to the study participants and back translated in English to the investigators. Key informant interview was used to collect qualitative data from four different sectors involved GBV survivors care pathways, namely: a health worker, a civil society worker, a law enforcer, and a community leader. Data were collected by the three primary investigators themselves.

### Data management and Analysis

The simple quantitative data was coded and entered in Kobo collect and later exported to STATA MP version 17 for further statistical analysis for bivariate and multivariable analysis to determine predictors of GBV among the participants.

### Quantitative Analysis

#### Descriptive Analysis

Categorical variables were summarized using frequency and percentage.

### Bivariate Analysis

Simple Poisson regression was used to assess associations between categorical variables, such as demographic characteristics and GBV among the participants. P<0.005 was considered statistically significant

### Multivariable Analysis

Multiple Poisson regression models was employed to identify predictors of GBV among the Participants for factors with p<0.2 in bivariate analysis. Adjusted prevalence ratios with corresponding 95% confidence intervals and p values were reported. P<0.05 was statistically significant.

### Qualitative Analysis

Qualitative data from FGDs underwent thematic analysis to identify key themes related to GBV among the participants. We use atlas ai. For the analysis.

## Results

### Characteristics of the participants

We enrolled 300 participants for this study, about half were in the age group 15 to 24 years (50.3%, n=151), education attain was primary (52%, n=165), married/ cohabiting (56.3%, n=169) and were in monogamous families (59.7%, n=179). More than two thirds were Dinka by tribe (67.3%, n=202). Majority were Christians (99.7%, n=299). Over a third had children ranging between 1 to 4 (39.3%, n=118), no child (35%, n=105) and were professionally employed (46.3%, n=139). **Table 1** summarizes the demographic characteristic of participants.

**Table 1:**
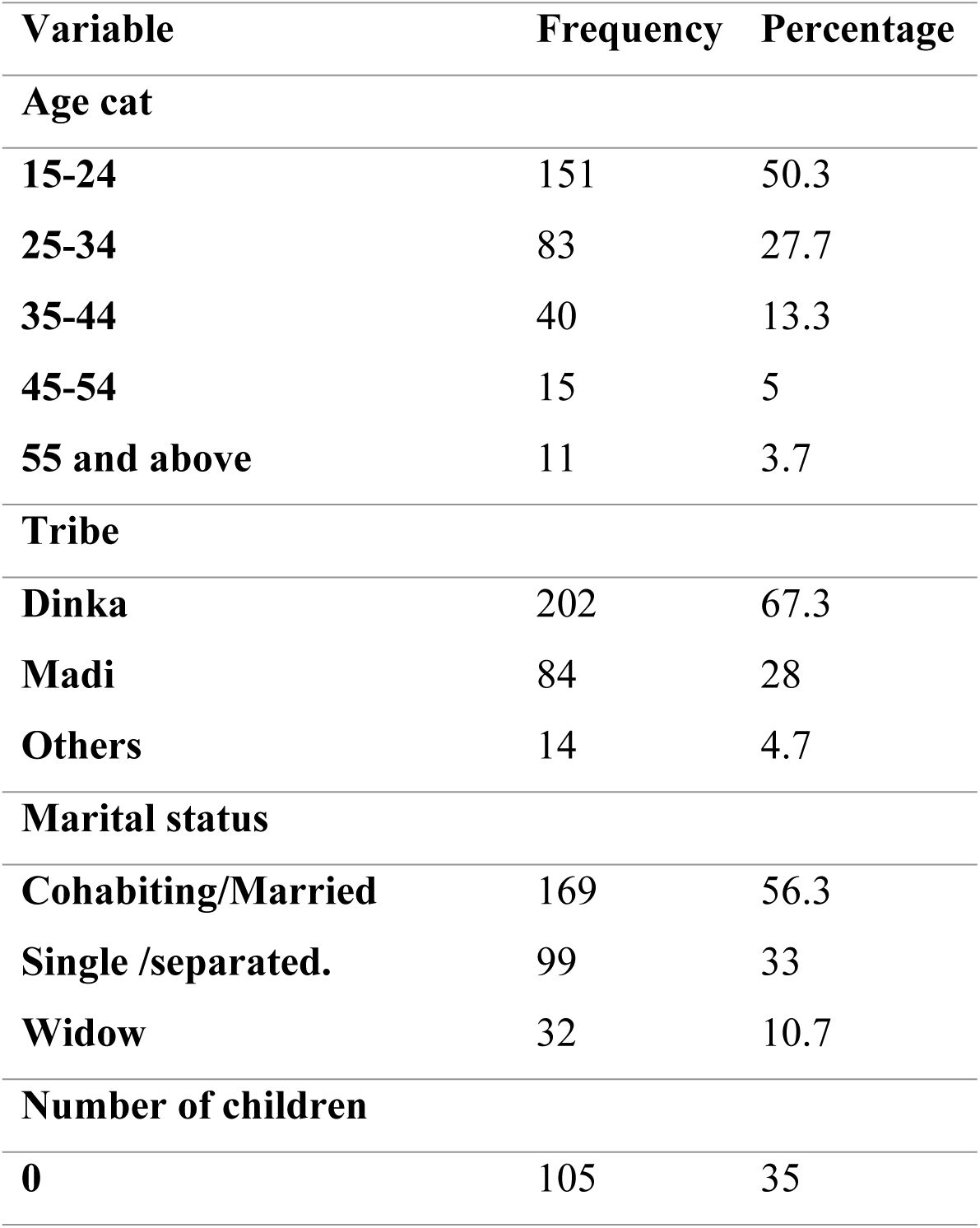
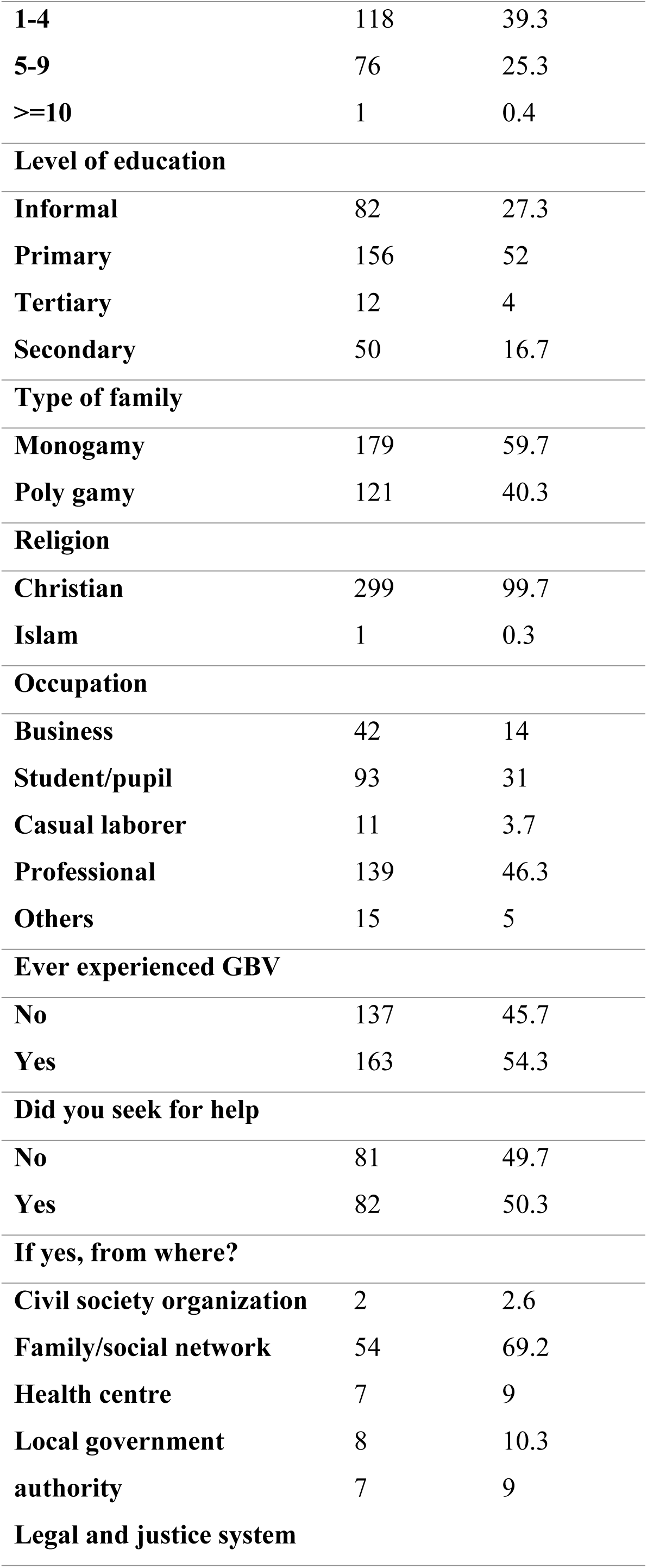
Characteristics of the participants.

| Variable | Frequency | Percentage |
| --- | --- | --- |
| **Age cat** |  |  |
| **15-24** | 151 | 50.3 |
| **25-34** | 83 | 27.7 |
| **35-44** | 40 | 13.3 |
| **45-54** | 15 | 5 |
| **55 and above** | 11 | 3.7 |
| **Tribe** |  |  |
| **Dinka** | 202 | 67.3 |
| **Madi** | 84 | 28 |
| **Others** | 14 | 4.7 |
| **Marital status** |  |  |
| **Cohabiting/Married** | 169 | 56.3 |
| **Single /separated.** | 99 | 33 |
| **Widow** | 32 | 10.7 |
| **Number of children** |  |  |
| **0** | 105 | 35 |
| **1-4** | 118 | 39.3 |
| **5-9** | 76 | 25.3 |
| **>=10** | 1 | 0.4 |
| **Level of education** |  |  |
| **Informal** | 82 | 27.3 |
| **Primary** | 156 | 52 |
| **Tertiary** | 12 | 4 |
| **Secondary** | 50 | 16.7 |
| **Type of family** |  |  |
| **Monogamy** | 179 | 59.7 |
| **Poly gamy** | 121 | 40.3 |
| **Religion** |  |  |
| **Christian** | 299 | 99.7 |
| **Islam** | 1 | 0.3 |
| **Occupation** |  |  |
| **Business** | 42 | 14 |
| **Student/pupil** | 93 | 31 |
| **Casual laborer** | 11 | 3.7 |
| **Professional** | 139 | 46.3 |
| **Others** | 15 | 5 |
| **Ever experienced GBV** |  |  |
| **No** | 137 | 45.7 |
| **Yes** | 163 | 54.3 |
| **Did you seek for help** |  |  |
| **No** | 81 | 49.7 |
| **Yes** | 82 | 50.3 |
| **If yes, from where?** |  |  |
| **Civil society organization** | 2 | 2.6 |
| **Family/social network** | 54 | 69.2 |
| **Health centre** | 7 | 9 |
| **Local government** | 8 | 10.3 |
| **authority** | 7 | 9 |
| **Legal and justice system** |  |  |

### Prevalence of GBV among the participants

More than half of the participants experienced GBV (54.3%, n=163) **Figure 1**. The most experienced GBV among the participants was emotional violence (23%, n=37) followed by physical (19%, n=31), multiple forms (23%, n=37), economical (17, n=28), other forms (4%, n=6) and sexual violence (2%, n=3) **Figure 2**.

**Figure 1:**
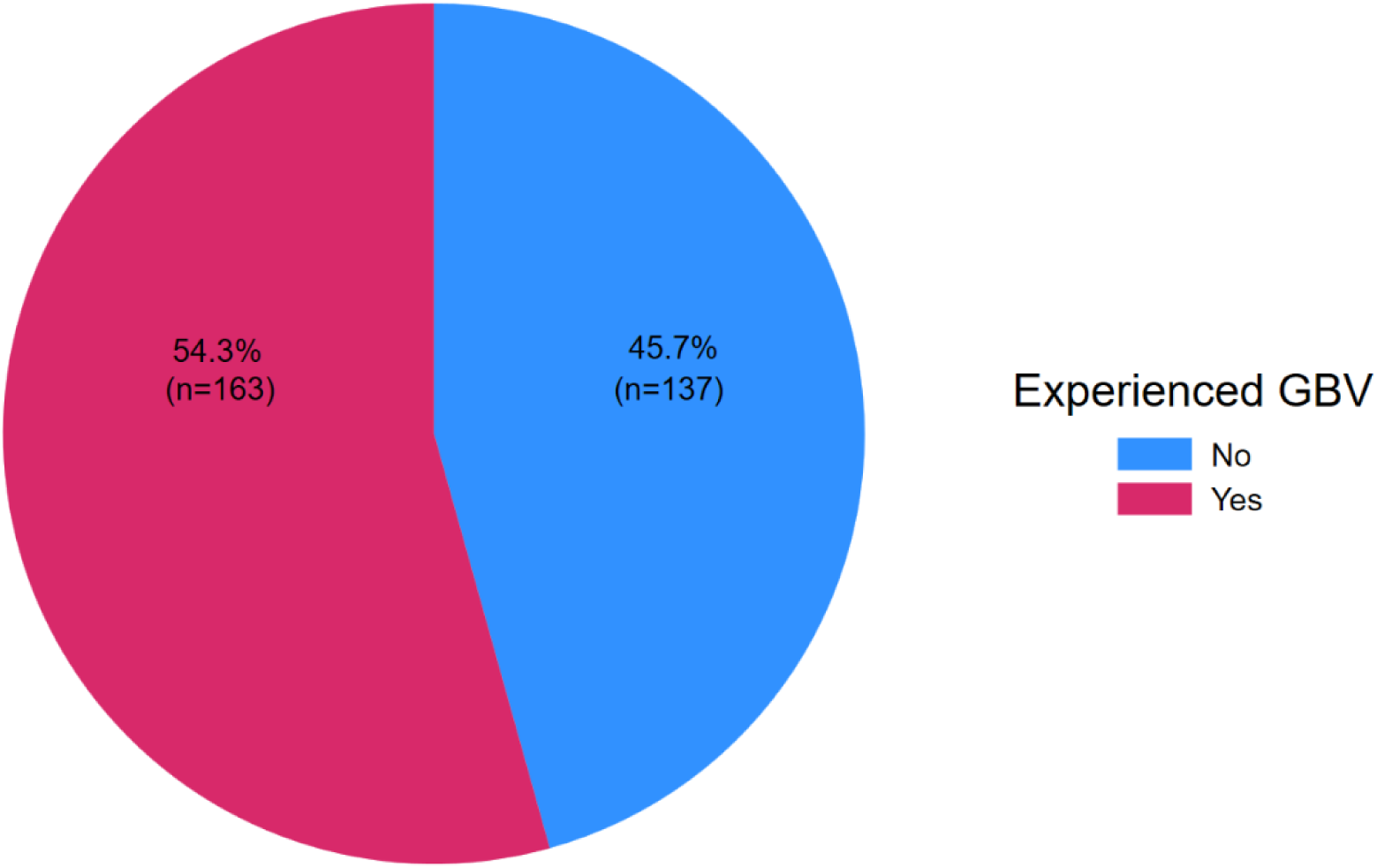
GBV among the participants.

**Figure 3:**
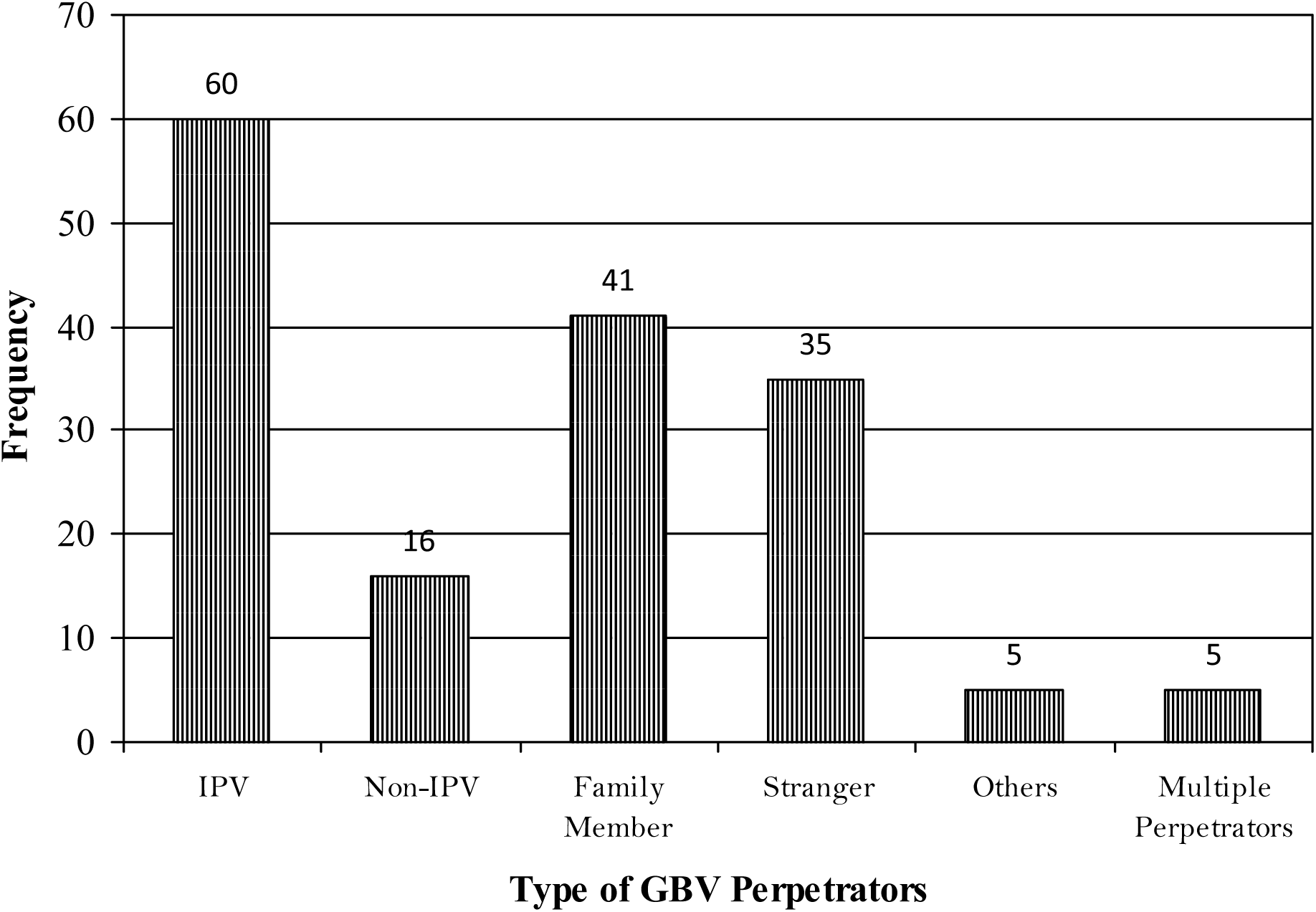
Perpetrators of GBV.

**Figure 4:**
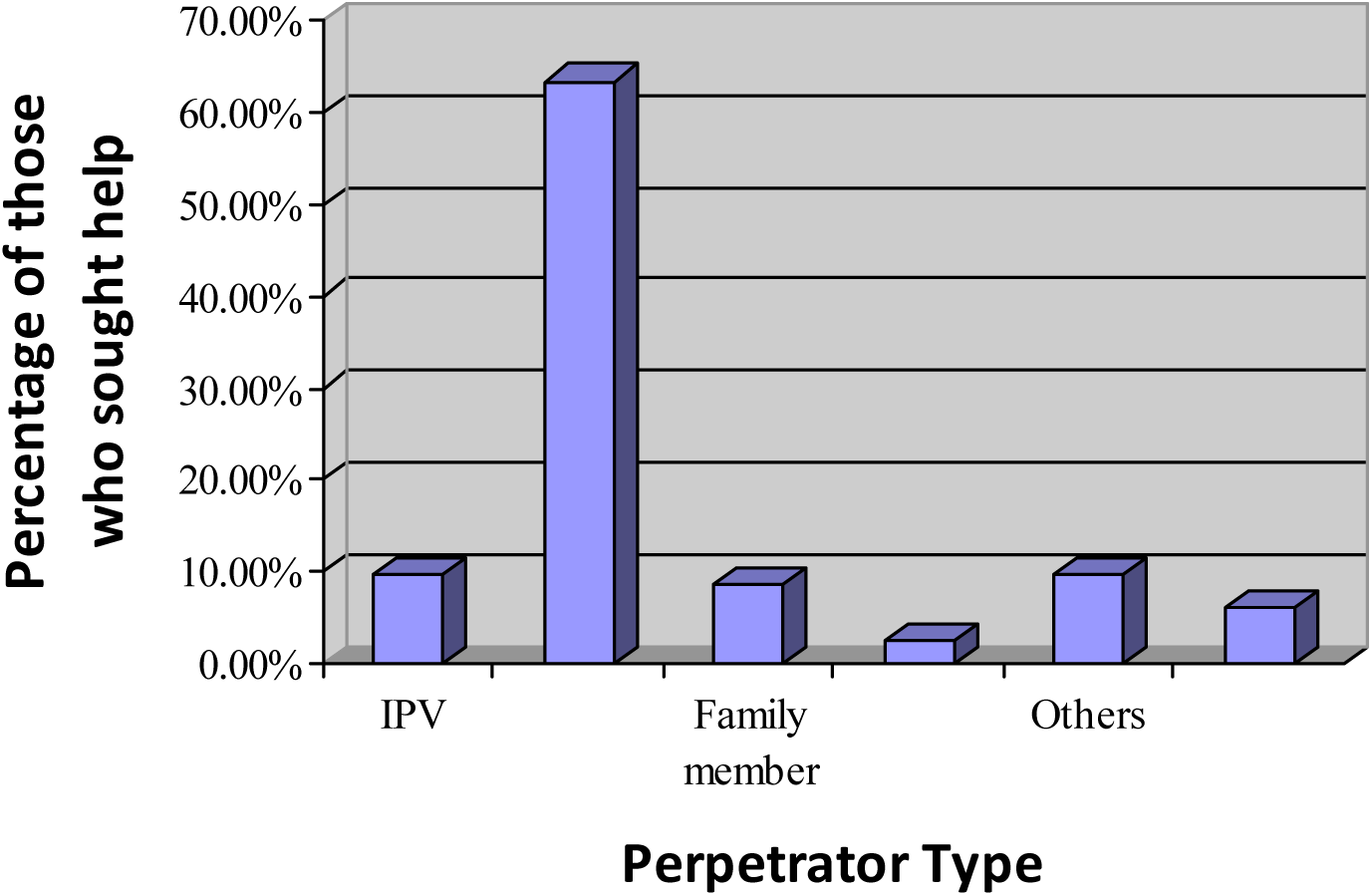
Bar graph showing prevalence of help seeking according to GBV perpetrator type. *Source: Primary Data*

### Factors associated with GBV among the participants

At Bivariate analysis, **Table 2**, Factors s with p<0.2 were taken for multivariable modified Poisson regression analysis **Table 3**. Factors independently associated with GBV among the participants were being Madi (Adjusted Prevalence rate: 1.6, 95% Confidence interval: 1.28-1.95, p<0.001) and others by tribe (Adjusted Prevalence rate: 1.7, 95% Confidence interval: 1.23-2.29, p=0.001), cohabiting/marriage (Adjusted Prevalence rate: 1.5, 95% Confidence interval: 1.12-2.04, p=0.007) and in polygamy family (Adjusted Prevalence rate: 1.3, 95% Confidence interval: 1.09-1.62, p=0.004).

**Table 2:** Bivariate analysis for factors significantly associated with GBV among the participants.

| Variable | All | GBV |  | Crude Prevalence | P value |
| --- | --- | --- | --- | --- | --- |
|  | (N=300) | No | Yes | Ratio (95% CI) |  |
|  | Frequency | (n=137) | (n=163) |  |  |
|  | (%) | Frequency (%) | Frequency (%) |  |  |
| Age cat |  |  |  |  |  |
| 15-24 | 151(50.3) | 76(55.5) | 75(46) | 1.8(0.68-4.85) | 0.231 |
| 25-34 | 83(27.7) | 31(22.6) | 52(31.9) | 2.3(0.86-8.13) | 0.097 |
| 35-44 | 40(13.3) | 13(9.5) | 27(16.6) | 2.5(0.92-6.66) | 0.073 |
| 45-54 | 15(5) | 9(6.57) | 6(3.7) | 1.5(0.46-4.63) | 0.513 |
| 55 and above | 11(3.7) | 8(5.8) | 3(1.8) | Reference |  |
| Tribe |  |  |  |  |  |
| Dinka | 202(27.3) | 108(78.8) | 94(57.7) | Reference |  |
| Madi | 84(28) | 26(19) | 58(35.6) | 1.5(1.21-1.82) | <0.001 |
| Others | 14(4.7) | 3(2.2) | 11(6.8) | 1.7(1.24-2.31) | 0.001 |
| Marital status |  |  |  |  |  |
| Cohabiting/Married | 169(56.3) | 65(47.5) | 104(63.8) | 1.4(1.08-1.78) | 0.011 |
| Single /separated. | 99(33) | 55(40.2) | 44(27) | Reference |  |
| Widow | 32(10.7) | 17(12.4) | 15(9.2) | 1.1(0.69-1.62) | 0.808 |
| Level of education |  |  |  |  |  |
| **Informal** | 82(27.3) | 35(25.6) | 47(28.8) | Reference |  |
| **Primary** | 156(52) | 71(51.8) | 85(52.2) | 0.9(0.67-1.36) | 0.7881 |
| **Tertiary** | 50(16.7) | 27(19.7) | 23(14.1) | 0.8(0.9-1.32) | 0.387 |
| **Secondary** | 12(4) | 4(2.9) | 8(4.9) | 1.2(0.55-2.6) | 0.693 |
| **Type of family** |  |  |  |  |  |
| **Monogamy** | 179(59.7) | 90(65.7) | 89(54.6) | Reference |  |
| **Poly gamy** | 121(40.3) | 47(34.3) | 74(45.4) | 1.1(0.90-1.67) | 0.188 |

**Table 3:** Regression Analysis for factors independently associated with GBV among the participants.

| **Variable** | **Adjusted Prevalence Ratio**<br>**(95% Confidence Interval)** | **P value** |
| --- | --- | --- |
| **Age cat** |  |  |
| **15-24** | 2.1(0.82-5.63) | 0.120 |
| **25-34** | 2(0.75-5.21) | 0.170 |
| **35-44** | 2.2(0.85-5.88) | 0.102 |
| **45-54** | 1.2(0.39-3.51) | 0.788 |
| **55 and above** | Reference |  |
| **Tribe** |  |  |
| **Dinka** | Refence |  |
| **Madi** | 1.6(1.28-1.95) | <0.001 |
| **Others** | 1.7(1.23-2.29) | 0.001 |
| **Marital status** |  |  |
| **Cohabiting/Married** | 1.5(1.12-2.04) | 0.007 |
| **Single /separated.** | Reference |  |
| **Widow** | 1.5(0.95-2.38) | 0.079 |
| **Type of family** |  |  |
| **Monogamy** | Reference |  |
| **Poly gamy** | 1.3(1.09-1.62) | 0.004 |

**Table 4:** Showing GBV help seeking association with GBV forms.

| **Forms of GBV** | **Occurrences** | **Percentage** |
| --- | --- | --- |
| **Physical violence** | 26 | 31.71 |
| **Sexual violence** | 2 | 2.44 |
| **Emotional and psychological violence** | 27 | 32.93 |
| **Harmful traditional practices** | 1 | 1.22 |
| **Socio-economic violence** | 8 | 9.76 |
| **Other forms** | 0 | 0.00 |
| **Multiple forms** | 18 | 21.94 |

### Prevalence of GBV according to the type of Perpetrators

About a third 60(37.04%) of GBV experienced were Intimate Partner Violence (IPV), 16(9.86%) non-IPV, 41(25.31%) from a family member, 36(22.22%) from strangers and 5(3.08%) from others. A very small number 5(3.08) experienced GBV from multiple perpetrators.

### Help Seeking

Of the participants who experienced GBV over half of them (50.3%, n=82) sought help from the different GBV referral pathway structures in the settlement. Of these more than two thirds (69.2%, n=54) sought help from the family/social network, with small percentages (10.3%, n=8., 9%, n=7., 9%, n=7. and 2.6%, n=2) from local government authority, health center, legal and justice system and civil society organizations respectively. while 81(49.70%) respondents did not seek redress, and one respondent did not reply to our question.

### Help Seeking according to GBV forms

Of those who sought help for GBV, 31.71(26 respondents) had experienced physical violence, 2.44% (2 respondents) had experienced sexual violence, 32.93% (27 respondents) had experienced Emotional & Psychological violence, 1.22% (1 respondent) had experienced Harmful traditional practices, 9.76% (8 respondents) had experienced Socio-economic violence, while there were no respondents who experienced other forms of violence who sought help, as shown in the table below.

### Help seeking according to the type of perpetrators

About two thirds 52(63.49%) of non-IPV sought help, only 10(9.76%) Victims of IPV sought help, 7(8.54%) victims of violence committed by family members sought help, 2(2.44%) victims of stranger related violence sought help, and 8(9.76%) of violence committed by others sought for help. 5(6.10%) of violence caused by both family members and stranger sought for help and no help was sought for violence committed by both intimate partner & family member.

### Where the Victim sought help from first

Of those who sought redress, over half 53(59.55%) sought from family/social network, 8(8.99%) went to the health centers, 13(14.60%) from local government authorities, 7(7.86%) from legal and justice, 3(3.34%) from civil society organization, and 5(5.62%) went to other authorities to seek redress as shown in the pie chart below. Of those who did not seek redress, 37(46.84%) were due to survivor related factors, and 33(41.77%) due to cited family, community and social network related factors. The rest had factors related to other sectors of the care pathway.

### Qualitative arm

Key informant interview covering the general overview of GBV in the settlement, the kind of services rendered, the statistics of GBV in their facility and the factors affecting help seeking revealed that:

GBV was a common occurrence in the settlement affecting mainly women. Commonly occurring was physical violence especially at water collection points due to struggle for water, hunger or food shortage and cultural practices. Other forms included cited by all the key informants included domestic violence, rape, forced and child marriages which are under reported but followed up by the different stakeholders. “The trend was dropping however there is a surge due to the COVID-19 lock down which led to reduction in the food ration/take home cash with associated economic difficulties,” remarked the RWC II.

On average, the health service provider, MTI, recorded 20-30 cases of physical violence and about 7 cases of SGBV monthly in their facilities. The RWC II received around 15 cases of GBV monthly mainly domestic violence, and few other forms like sexual violence. Ayilo police station, recorded 9 or more cases of physical violence of monthly, has reducing cases of rape and forced abortion and no data on other forms of GBV.

Available services included GBV minimal health care package for survivors offered by MTI; legal representation offered by Refugee Law Council; Law enforcement offered by Uganda Police; Psychosocial support by TPO; Follow-up and rehabilitation by Lutheran World Federations (LWF). Office of the Prime Minister (OPM) coordinates and oversees the operations of all these partner organizations. The local leaders mainly do sensitization, reporting of perpetrators, peace mediation and dialogue at family level. There is inter-sectoral linkages and referrals. However, service utilization is low at some sectors.

The common factors affecting help seeking cited by the key informants included but are not limited to; stigma and fear of being exposed, acceptability by culture and social norms of some violence like forced and child marriage, lack of confidentiality from service providers, consequences of reporting like arrest and/or murder of the perpetrator and influence of local leaders on most cases. Less common factors included young victims being unable to report cases themselves, no place of custody for GBV victims/perpetrators, knowledge gap and inadequate personnel to offer relevant intervention at community level.

## Discussions

The study found out that the prevalence of GBV was 54%. This is consistent with that reported by UDHS, 2016 (56%) and 51% according to the 2017 global gender-based gap report in Uganda thus implying no change over the years (12). This could be higher than found because of under-reporting and failure to report or talk about GBV especially among the Dinka tribe (Key Informant Interviews report (KII)). These findings conform with that of the World Bank Group 2019 that GBV is worsened in emergency situations where social support structures have broken down (13). However, this prevalence is much higher than outcomes of systematic review conducted by Vu etal on prevalenve of seuxal and gender based violence among female refugees in complex humanitarian emergencies which found it at 21.4% (14). Possible reason for this difference is that this study was a systematic review incorporating 19 studies with varied results and resultant. In all those studies, most investigators concluded possibilities of low level of declarations among participants with possibilities of higher prevalence. Kinyanda and his colleagues found prevalence of IPV at 43,7% among women in post conflict Eastern Uganda (15), while Wandera and his colleagues focused on Intimate Partner Sexual Violence (IPSV) among married women in Uganda and found prevalence at 17% (16).

Our study found out prevalence of GBV highest among women less than 24 years 45.68% with total of 77.7% of those who experience GBV being below 34 years of age. This finding is consistent with other studies done in Uganda (15–17) and those done outside Uganda (18). This could be because they are less experienced and less knowledgeable about the risk factors leading to GBV hence easy for the perpetrators to manipulate them. A law enforcement officer, one of our key informants, also quoted that GBV is on the rise among people of concern in the settlements because of inability to guide and control the young people.

Our study found that being from madi tribe and other tribes other than dinka tribe had significant asssociation with GBV ((adjusted prevalence rate 1.6, 95% CI.: 1.28-1.95, p<0.001) and (adjusted prevalence rate 1.7, 95% CI: 1.233-2.29 p=0.001) respectively) but this could also be explained by the openess by these tribes in discussing about GBV. The Dinka tribe could have mastered the Ugandan law hence manipulate it to suit their tradition without being apprehended “For instance, child marriages are settled from South Sudan and couple just return to the settlement after, and thus dodging the Ugandan laws on GBV,” said one of the key informants. Furthermore, the concealed nature of GBV overshadows the extent and reality of its existence as reported by Nkiriyehe et al (19) and Opiro (20). A refugee setting, especially in this context is characterized by factors such as child headed families or being parentless confirmed by the proportion of women whose partners were staying in South Sudan. This agrees with the (21) report which highlighted that the majority of people affected by humanitarian crisis are women and children.

Relating marital status, just like in Wandera et al, 2015 (16), the married and cohabiting had significant association with GBV (adjusted prevalence rate 1.5%, 95% CI, 1.12-2.02, p=0.007). This this is consistent with the findings of this study that the biggest numbers of perpetrators at a prevalence of 37% were intimate partners. Global and regional estimates of violence against women, UNHCR had similar findings of intimate partners being the most perpetrators (22).

The prevalence of GBV is higher amongst those with no (28.8%, n=47) or low level of education (52.2%, n=85.0%) and low socio-economic status though this was not statistically significant. Lack of knowledge or information that comes with exposure to education makes them more susceptible than those with higher level of education. It can also be due to probably strongly believe and adherence to cultural norms or being oppressed easily (23), and high level of economic dependence (>80.00%) with possible socio-economic discrimination as noted by Beth et al (24). This was evidenced by high percentage of those with informal education (27.30%, n=82) and majority having attained only primary level of education (52.0%, n=156). More than half of the respondents were dependent on UNHCR handout only with some receiving support from family and friends back home (25).

There was minimal variability in the prevalence of GBV according to religion and family type. 98% of the respondents were Christians and majority had monogamous family. However, polygamy which was significantly associated with GBV in our multivariate analysis (adjusted Prevalence rate 1.3, 95% CI:1.09-1.62, p=0.004) was more common among the Dinka and could have somehow added to the higher prevalence among this tribe though not statistically significant as demonstrated in study by Bisika (26). Other plausible explanation for polygamy contributing to GBV is the extra economic stress the family head has in providing basic needs for the multiple wives and their children. Temporary separation especially among the Dinka culture, whereby a woman leaves the husband when the baby is around 3 months to stay with her parents and only returns when the child is 2 years (KII result) or distance due to other reason for instance husband still studying contributed to the higher prevalence of emotional/psychological and socio-economic violence.

The majority (35%) of the respondents experienced emotional and psychological violence, (19%) physical violence, 17% socio-economic violence and only 2% and 1% experienced sexual and harmful traditional practices respectively. Hence emotional and psychological violence are highly prevalent in this community compared to physical and sexual violence which are lower compared with the report that 1 in 3 women have been reported to have experienced physical and sexual violence (27). Most probable factors included low socio-economic status and deprivation of care and attention from their spouses because of their continued absence. Physical violence is most likely due to volatile emotions following continued frustrating situations such as lack of basic needs, unemployment, and separation from families among others.

SGBV in particular was low 2% (3) which contrasts a study done in post violence northern Uganda which was at 47% (20), war related sexual violence in Kitgum 28.6% (28) and IPSV among married women in Uganda (16). This is probably due to under reporting or because the Dinka community is reserved at sharing it as pointed out by the Key Informants. Contrary to this finding, Vu et al 2014 found that 1 in 5 female refugees had experienced SGBV (14) and that the number could be even higher due to hesitancy at disclosure (29).

Many experienced multiple forms of GBV with related causes or risk factors mainly from intimate partners (37.00%) and the married/ co-habiting suffered more (83.9%), a similar finding with WHO reports (27).

### Help Seeking

Of the 162 survivors, only 83 sought redress. A quarter (25.61%) of the respondents who experienced gender-based violence and sought help were single, 52.44% were married, 2.44% was divorced while 7.32% were cohabiting and 12.2% were widows. This is contrary to who cited that married women hardly sought redress after experiencing GBV (30). This comes with one knowing their rights and a GBV care system that protects sensitive survivors such as the married ones from the perpetrator and the community at large.

The elderly and those with large family size more than 10 children had low help seeking compared to the young with smaller family size as they relied on traditional informal sources. This is consistent with findings by McCleary-Sills in his survey in Tanzania whereby the younger sought help but from the family and social network not from the formal settings (30).

There was proportional reporting with respect to tribe, occupation, religion, and family type. However, help seeking was high among victims of socio-economic, emotional and psychological violence followed by physical violence and least with SGBV agreeing with Undie, Birungi and Jane who also found that only 10% disclose SGBV(29,31). Violence committed by non-partners and strangers were reported as opposed to those done by intimate partner and/or family member and concurs with the finding of (32). Possible explanation for this is the fact that intimate partners have more reasons to forgive and reconcile and also the fact the perpetrators could be the bread winners in the family compared to strangers (33).

Help seeking was mainly from the family/social networks (63%) which don’t offer necessary care package in agreement with Mc Cleary - Sills, 2013 and United Nations Economic and Social Affairs, 2015 (30)(34). About 10% went to the health facilities and local government authorities respectively, 9% went to legal and justice, the least number sought redress from civil society organization and/or police. This agrees with (32) where only 14% and 13% reported cases of IPV and non-IPV to police and UN social and economic affairs report.

Other services remain under-utilized despite their establishment within the settlement contrary to (21). Absurdly and not unique to only refugee settings, GBV is widely not reported and consequently survivors don’t seek redress. Commonly, the family of the survivor and that of the perpetrator reach favorable agreements not considering the survivor which equally leads to grave social consequences as reported by McCleary – Sills, 2013 (30).

### Factors affecting help seeking

Help-seeking refers to the methods that victims of GBV get support from either formal like hospitals and legal help or informal systems from elders, friends among others (35). Besides the families of the victim and the perpetrator settling cases at family level, the major reason people don’t seek redress is stigma and fear of being exposed. This was also reported by the camp commandant and the Police officer reported in the key informants’ interview (KII). Survivor related factors (47.00%) included shame, guilt, fear, discrimination while family, community and social network related factors included failure to get a suitor for marriage, fear of losing the family bread winner, consequences of going against elders and tradition, family discrimination and threat of violence. Some of these findings are consistent with what were also reported by Tin and Subadra Panchanadeswaran in 2009 and Rose Mogga (36,37). In a more recent study by Melgar et al found fear for retaliation, though not reported as a major factor hindering help seeking in our study was a major factor with up to 40% of people didn’t get help due to fear for retaliation (38). Many other studies in other settings have also shown clearly that stigmatization is one of the major factors that victims fear and so not seek for help after falling victims of gender based violence (39,40). Other studies done in South Sudan demonstrated sociodemographic factors as well as where to seek for help were some of the factors that influenced the victims whether or not a victim will seek for help (41). Recent studies in the United States though completely different settings compared to ours found out that information gaps and communication struggles were the major factors hindering victims of IPSV from seeking for help (2).

Similar findings regarding factors affecting health seeking were given by the Key Informants and included stigma and fear of being exposed, suppression by culture and cultural practices especially bride price, lack of confidentiality from the people in the care pathway, consequences of reporting (risk of being murdered by community members and fear of arrest of either intimate partner or family member) and influence of local leaders on most cases. The young victims are unable to report cases themselves and there are no places of custody for GBV victims/survivors. Some sectors had knowledge gap and inadequate personnel to offer relevant intervention at community level.

Other forms of SGBV, for instance, early/forced marriage is socially acceptable within the community’s social and cultural norms. The Dinka community assumes that a girl is mature and ready for marriage as soon as she receives her menarche. She is advertised to men and whoever bids higher number of cattle takes her irrespective of her choice or interest. This was also reported by Koenig in 2013(42).

### Conclusions

More females had experienced GBV compared to those who did not. The prevalence was at 54% though there is a likelihood it can be higher than this for factors cited in the discussion. Emotional and physical was the greatest form of GBV experienced. The main perpetrators were intimate partners and family members. The risk factors were younger age, unemployment, poverty, no/low level of education and or high level of dependence. Precipitating factors include cultural practices and beliefs, influence of local leaders on GBV cases, lack of enough safe spaces and no places of custody for GBV victims.

Much as GBV services are offered at no cost at the refugee camp, only 51% of the survivors sought redress and majority of them sought from family/social network only which doesn’t provide a minimal care GBV package. The major reasons people did not seek redress included stigma and fear of being exposed, suppression by culture and cultural practices especially bride price, lack of confidentiality from the people in the care pathway, bad consequences of reporting, young victims being unable to report cases themselves, no place of custody for GBV victims, knowledge gaps and inadequate psychosocial and socio-economic support at the community level.

## Ethical approval and consent to participate

The ethical clearance to execute the study was obtained from Gulu University Research Ethics Committee and was taken to Office of the Prime Minister Refugee desk in Kampala. Permission was granted in writing and letter copied to the Refugees desk Adjumani and the Assistant camp commandant Ayilo Refugee settlement for further permission to access the settlement. Clear ground rules were issued and were adhered to strictly. A written consent and ascent, or thumb printing was obtained from study subjects and/or parents/guardians who were willing to participate in the study. The interview was conducted with maximum privacy and confidentiality maintained during and after the interview. The questionnaire was approved by the ethical committee and was anonymously and directly administered by the investigators. The participants had the rights to withdraw at any point from the interview, in case they felt uncomfortable to continue. Those who were re-traumatized during the interview were linked to the relevant stakeholders like health facility for further management.

## Consent for Publication

All the authors have consented and Gulu University as an institution encourages publication of important findings to make it available to the rest of scholars across the globe. Consent for publication by study participants are included in the consent to participate in the study.

## Availability of data and materials

Yes

## Authors contributions

Denis Odongo, Aeku Job Michael, Ocora Boniface and Okot Jerome generated the topic, developed the proposal, collected data, analyzed the data, and wrote this manuscript all through with guidance and contributions from their supervisor Keneth Opiro and Pebalo Francis Pebolo.

## Data Availability

Data is available upon reasonable request

No

## Acknowledgements

We acknowledge the Almighty God for the gift of life, health, and knowledge for developing this piece of work. We are very thankful to the researchers whom we cited their publications in our work and all the different stakeholders who made the study a success i.e. the camp leadership of Ayilo settlement, implementing partners such as Medical Teams International, Lutheran World Federations among others and Office of the Prime Minister. We are greatly indebted to our supervisors Dr. Kenneth OPIRO and Dr. Pebalo Francis Pebolo whose efforts, patience, assistance and guidance made this work a success as well as assisting in writing this final manuscript for publication. Lastly, we thank our families and friends for their prayers, encouragement, and support.

